# Overestimation of School-Based Deworming Coverage Resulting from School-Based Reporting

**DOI:** 10.1101/2022.04.11.22273689

**Authors:** William Sheahan, Roy Anderson, Kumudha Aruldas, Euripide Avokpaho, Sean Galagan, Jeanne Goodman, Parfait Houngbegnon, Gideon John Israel, Venkateshprabhu Janagaraj, Saravanakumar Puthupalayam Kaliappan, Arianna Rubin Means, Chloe Morozoff, Emily Pearman, Rohan Michael Ramesh, Amy Roll, Alex Schaefer, Sitara S. R. Ajjampur, Robin Bailey, Moudachirou Ibikounlé, Khumbo Kalua, Adrian J.F. Luty, Rachel Pullan, Judd L. Walson, Kristjana Hrönn Ásbjörnsdóttir

## Abstract

**Background:** Soil Transmitted Helminths (STH) infect over 1.5 billion people globally and are associated with anemia and stunting, resulting in an annual toll of 1.9 million Disability-Adjusted Life Years (DALYs). School-based deworming (SBD), via mass drug administration (MDA) campaigns with albendazole or mebendazole, has been recommended by the World Health Organization to reduce levels of morbidity due to STH in endemic areas. DeWorm3 is a cluster-randomized trial, conducted in three study sites in Benin, India, and Malawi, designed to assess the feasibility of interrupting STH transmission with community-wide MDA as a potential strategy to replace SBD. This analysis examines data from the DeWorm3 trial to quantify discrepancies between school-level reporting of SBD and gold standard individual-level survey reporting of SBD.

**Methodology/Principal Findings:** Population-weighted averages of school-level SBD calculated at the cluster level were compared to aggregated individual-level SBD estimates to produce a Mean Squared Error (MSE) estimate for each study site. In order to estimate individual-level SBD coverage, these MSE values were applied to SBD estimates from the control arm of the DeWorm3 trial, where only school-level reporting of SBD coverage had been collected.

In each study site, SBD coverage in the school-level datasets was substantially higher than that obtained from individual-level datasets, indicating possible overestimation of school-level SBD coverage. When applying observed MSE to project expected coverages in the control arm, SBD coverage dropped from 89.1% to 70.5% (p-value < 0.001) in Benin, from 97.7% to 84.5% (p-value < 0.001) in India, and from 41.5% to 37.5% (p-value < 0.001) in Malawi.

**Conclusions/Significance:** These estimates indicate that school-level SBD reporting is likely to significantly overestimate program coverage. These findings suggest that current SBD coverage estimates derived from school-based program data may substantially overestimate true pediatric deworming coverage within targeted communities.

## Introduction

Soil-transmitted helminths (STH) are a group of parasites (*Ascaris lumbricoides, Ancylostoma duodenale, Necator americanus, and Trichuris trichiura*) estimated to infect over 1.5 billion people globally (1). While these infections do not directly result in significant mortality, they do contribute substantially to morbidity, causing malnutrition, anemia, and stunting, resulting in a toll of 1.9 million Disability-Adjusted Life Years (DALYs) in 2019 (2). To mitigate the impact of these parasitic infections, the World Health Organization (WHO) has endorsed a strategy of deworming via mass drug administration (MDA) with albendazole or mebendazole for pre-school-age children (PSAC) and school-age children (SAC), women of childbearing age, and adults in high-risk occupations, including agricultural labor and mining. Deworming campaigns in 2018 treated over 676 million SAC, representing approximately 53% of all children estimated to be at risk for infection (1).

Measurement of program coverage is a critical aspect for deworming campaigns and is the primary metric by which WHO sets programmatic targets. WHO guidance on conducting coverage surveys for preventative chemotherapy defines coverage as answering the fundamental question “how many people in need of treatment swallowed the drugs (3).” This guidance also notes that difficulty in determining the underlying population denominator of SAC in a program area may impede the accurate measurement of program coverage. School attendance may vary within program areas, and school enrollment data may not accurately reflect the number of students that attend school on the day of deworming treatment. Additionally, while many MDA campaigns for STH are conducted via school-based drug distribution, school-based MDA may not reach all at-risk SAC, especially those who do not attend school. To address this gap and protect against re-infection of SAC by disease reservoirs in the broader community, a number of studies have examined the effect of conducting community-wide MDA together with school-based MDA (4).

The DeWorm3 trial is a cluster-randomized trial examining the feasibility of interrupting the transmission of STH by comparing community-wide MDA with the standard of care school-based MDA in endemic areas of Benin, India, and Malawi (5). The analyses presented here use DeWorm3 trial data to examine the degree to which discrepancies in school-level reporting of SBD may lead to overestimation of SBD coverage when compared with individual-level reporting of SBD. Coverage estimates derived from these two sources of data may differ, even in overlapping populations. School-level MDA data may not reflect the same coverage levels as those from the individual-level MDA treatment records provided by drug distribution data, as school-level coverage estimates might be based on a denominator that includes only a subset of children who attend school on deworming days, potentially excluding SAC who do not attend school on those days, or, as is the case in India, children aged 17 or 18 years who have entered college and would be excluded from school-registry denominators. In Benin, children aged 5-14 years who were not enrolled in school were invited to go to their nearest school the day of MDA to be treated, but only those enrolled in school were included in school-registry denominators. Previous studies have suggested that such heterogeneous data reporting practices by schools during SBD may result in overestimates of deworming coverage when compared to individual-level data collection (6).

DeWorm3 provides a novel opportunity to analyze these SAC coverage data due to the rigorous and systematic conduct of annual household censuses that link the school each child attends to their physical home address. This allows for an improved understanding of the geographic catchment areas of each school in the study areas, as well as the underlying population denominators of each study cluster. These data are less commonly collected by programs or by researchers, who often only collect school-level deworming data. As a result, this analysis offers an opportunity to assess overestimation in traditional school-based coverage estimates. This comparison of school-level data to individual-level reporting has broad implications for understanding discrepancies in reported coverage for school-based MDA programs.

## Methods

### Study Design

All data used in this analysis come from the DeWorm3 trial. A detailed description of the trial methods have been reported elsewhere (5). The analysis in this paper is descriptive in nature, evaluating past results reported by the DeWorm3 trial by calculating new cluster-level coverage estimates to compare with previously reported estimates.

### Study Setting

The DeWorm3 trial operates in three primary locations. In Benin, the Institut de Recherche Clinique du Bénin and the French Government’s Institut de Recherche pour le Développement collaborate with the Ministry of Health to administer the trial in the commune of Come. In Malawi, trial operations are conducted by the Blantyre Institute for Community Outreach, the London School of Hygiene and Tropical Medicine, and the Ministries of Health and Education in the district of Mangochi. In India, trial operations are conducted in collaboration with the state government of Tamil Nadu by the Christian Medical College, Vellore (5). Each country included an entire administrative area (two administrative areas in India) inclusive of more than 80,000 individuals in each country site. These administrative areas were divided into 40 clusters, each containing a minimum of 1,650 individuals. Twenty of these clusters were randomized to receive community-wide MDA, while twenty receive standard of care SBD.

### Ethics statement

The DeWorm3 trial has been reviewed and approved by the Institut de Recherche Clinique au Bénin (IRCB) through the National Ethics Committee for Health Research (002-2017/CNERS-MS) of the Ministry of Health in Benin, The London School of Hygiene and Tropical Medicine (12013), The College of Medicine Research Ethics Committee (P.04/17/2161) in Malawi and the Christian Medical College Institutional Review Board in Vellore, India (10392). The overall study was approved by The Human Subjects Division at the University of Washington (STUDY00000180) and registered at ClinicalTrials.gov (NCT03014167). This particular analysis was also reviewed and approved by the Human Subjects Division at the University of Washington (Study 00012268).

### Consent procedures

Data collectors obtained consent / assent as appropriate prior to all data collection activities. Written informed consent was sought if the individual could write, while oral consent was given in the presence of a witness and documented with a thumbprint for any individual who could not write.

### Study Population

As described by Ásbjörnsdóttir et al., 2018, the DeWorm3 trial tests the feasibility of interrupting STH transmission throughout selected communities, using community-wide mass drug distribution for all age groups in the intervention clusters. However, this analysis maintains as its focus the treatment of school-age children (SAC) 5-14 years of age in Benin, 5-19 years of age in India, and 2-19 years of age in Malawi, who are treated at school in both arms of the trial. A baseline census conducted before the start of MDA in these three sites has been previously reported (7) in which all individuals from the households within each study area were enumerated and socio-demographic characteristics collected.

### Data Collection

Data collected through the DeWorm3 trial were used for this analysis. Data collection methods are detailed extensively in the original cluster-randomized trial protocol (6). The DeWorm3 trial has provided door-to-door community-wide MDA for STHs in intervention arm clusters, timed to follow the standard of care school-based MDA, while control clusters continue to receive standard of care school-based MDA only. Community-wide coverage surveys assessing self-reported coverage and treatment uptake in a subset of households in both arms are available. However, while individual-level drug distribution data are available in the intervention arm, control arm data are limited to routine school-level coverage data.

### Data Analysis

Data were analyzed using R Studio 3.6.2 (8) and QGIS 3.10.7 A Coruña (9), primarily leveraging the tidyverse package in R (10). Individual-level SBD coverage estimates were summarized to the cluster level based on the location of each respondent’s home. Because SAC are known to attend schools in clusters that are different from the clusters where they reside, no single school-level coverage estimate could be assumed to be directly representative of SBD coverage in the clusters in which the schools were located. School-level coverage estimates were summarized to the cluster level using a weighted average calculation. In this calculation, the reported coverage levels for all schools attended by SAC in each cluster were weighted by multiplying them by the proportion of SAC in the cluster who reported attending that school. For example, if there were 100 SAC in hypothetical cluster X, and thirty of them self-reported as attending school A, fifty reported themselves attending school B, and twenty reported themselves attending school C, the weights for schools A, B, and C, would be 0.3, 0.5, and 0.2 respectively.

This analysis leveraged the intervention clusters in which both individual-level and school-level SAC coverage estimates are available to quantify the discrepancy between these data sources. The individual-level data from community-wide MDA specifically identifies which SAC reported having been treated at school during the most recent round of school-based MDA. If we treat the individual-level estimates of previous coverage in each cluster as the reference value that acts as a gold-standard or “true” value, then over or under-estimation in the cluster-level coverage estimates from the school-level data can be measured by calculating the error, or the difference between those estimates and the gold-standard values from the individual-level data.

The difference between these “true” cluster-level estimates and the cluster-level estimates derived from the school-level data were calculated by subtracting each school-level estimate from each corresponding individual-level estimate and squaring that value. These residual errors were then added together and divided by the number of clusters to get an estimate of the mean squared error (MSE) of the combined estimates for each study site. This statistic is available from the following equation:

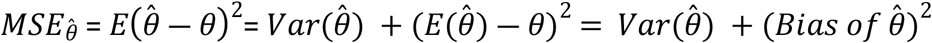

Where 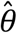 is the estimator of the unknown parameter *θ* (11), here taken to be the true SAC coverage at the cluster-level which is approximated by individual-level coverage estimates from the intervention arm.

After calculating the error in existing school-level SBD coverage estimates from the intervention arm, the overall site errors were applied to control clusters where only school-level SBD estimates were available. Multiplying the existing school-level SBD estimates by the ascertained error and subtracting this from the school-level SBD estimate allowed us to project the likely individual-level SBD coverage estimates in the control arm of the trial. Statistical significance for the difference in mean SBD coverage estimates was produced by paired t-test (12) for Benin and Malawi where the cluster means were normally distributed, and by Wilcoxon-Signed-Rank (13) test in India where the cluster means were distributed significantly different from normal according to a Shapiro-Wilk normality test (14).

### Catchment Area Analysis

The catchment areas of each school in the DeWorm3 registry were calculated to provide a more detailed understanding of where children within the study sites go for their SBD treatments. For each study site, the locations of every school in the trial’s school registry were mapped in QGIS, along with the residential locations of each child for whom self-reported school attendance was available.

A key component of this analysis included the integration of free text fields from surveys that capture school attendance for SAC treated in the community. While for the majority of SAC, their designated school was chosen from dropdown menus and can be easily matched to the school they attend, many respondents gave the names of schools that were not included in the trial’s predetermined school registries, or used alternative spellings, or colloquial names for their schools. These free-text responses were matched to their official school names in the registries through a process of visual fuzzy matching to identify common misspellings or translations, as well as quality control checks sent to study sites for confirmation of assumptions on close spelling matches. Between 10 and 17% of free-text entries for each site remained unmatched and were excluded from the catchment area creation process (Table 1).

**Table 1.** Proportion of School-Age Children in DeWorm3 Catchment Areas with Unmatched School Names, by Study Site.

|  | Matched (n) | Unmatched (n) | Total (n) | Proportion Unmatched |
| --- | --- | --- | --- | --- |
| Benin | 18140 | 3700 | 21840 | 0.17 |
| India | 21579 | 3929 | 25508 | 0.15 |
| Malawi | 38304 | 4246 | 42550 | 0.10 |

To obtain an initial understanding of the geographic range attributed to each school’s catchment area, the QGIS processing toolbox function “Concave Hull (k-nearest neighbor)” was utilized to create unique minimum-bounding geographies that included the area within which SAC reported attending each school. A naïve version of this analysis was conducted using all SAC whose responses were matched to a specific school.

In order to obtain more discriminatory catchment areas for use in further analysis, the catchment area creation process was repeated after removing statistical outliers by furthest geographical distance between SAC and their self-reported schools. Using the QGIS processing toolbox function “Join by Lines (hub lines),” the Euclidean distance was calculated between each SAC in the study area and the location of their self-reported school. In order to remove geographic outliers, the distribution of the distance variables was calculated separately for each study site, with histogram plots showing an approximately log-normal distribution in each case (Fig 1).

**Fig 1.**
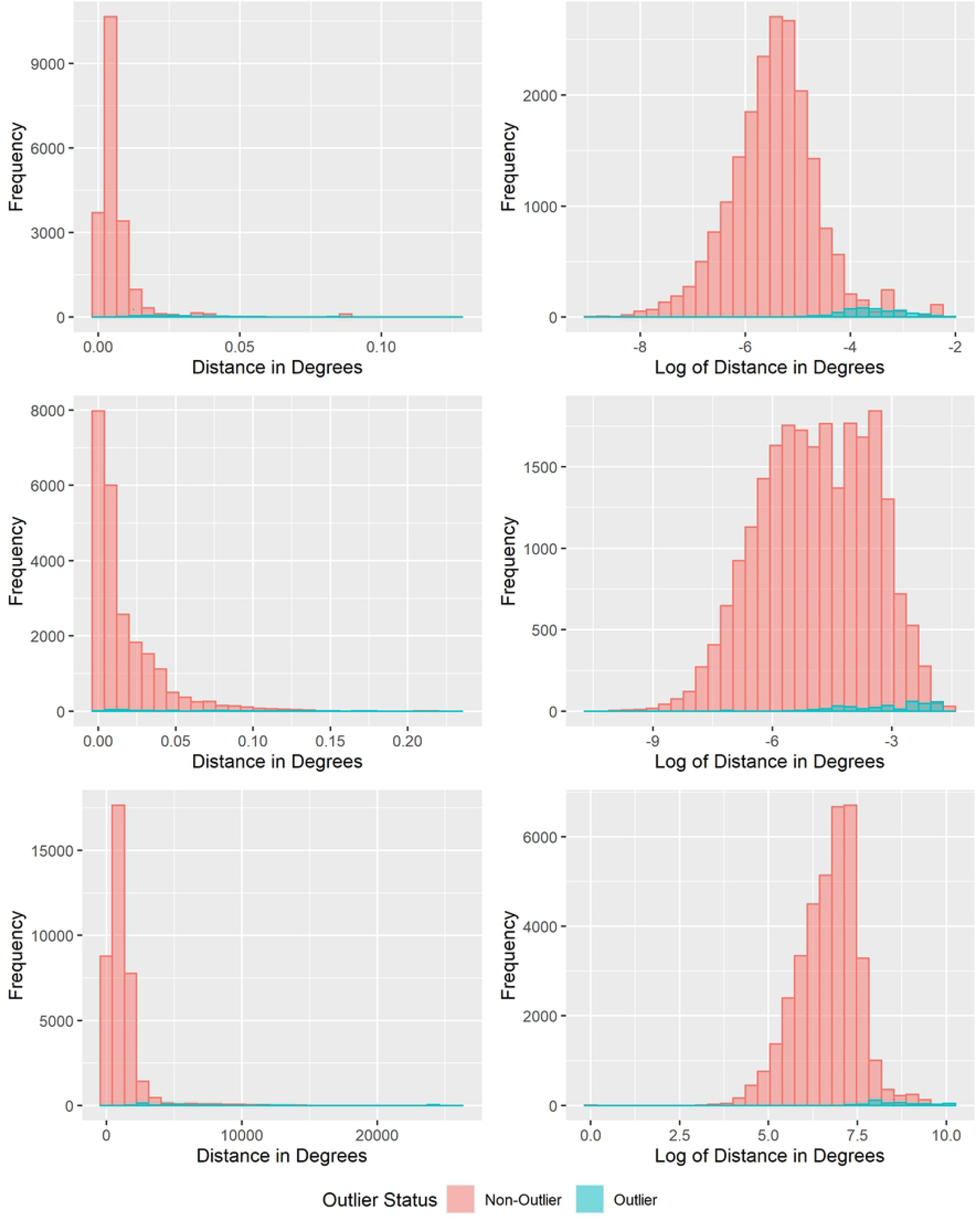
Histograms of the distance between School Age Children (SAC) and their self-reported schools. (A) Benin. (B) India. (C) Malawi. The left graph in each panel shows the distribution of the untransformed variable with distance measured in map units, on the right is the log-transformed variable.

Because of the approximately log-normal distribution of the distance between the SAC households and their reported schools, geographic outliers were removed for each school if the log-transformed distance between a respondent and their self-reported school was greater than the product of 1.96 times the log of the standard deviation for each school plus the log of the mean distance between each school and the SAC who reported attending them.

### Parent Study Power

No new sample size calculations were conducted for this analysis, as the requisite sample size powered to detect differences in final prevalence of STH infection between the control and intervention arms was calculated by the DeWorm3 trial team during the course of previously published analyses (5). These calculations resulted in a minimum site sample size of 80,000 individuals, with individual clusters containing a minimum of 1,650 individuals.

## Results

Cluster-level SAC SBD coverage was calculated using individual-level data from the clusters randomized to receive the community-wide MDA intervention annually in Malawi and Benin, and bi-annually in India. These estimates specifically determined the cluster-level coverage as the proportion of SAC reached during community-wide MDA who reported being treated during previous SBD. These SBD coverages were consistently lower than the WHO 75% coverage goal for SAC and PSAC (15), with lower than 50% SBD coverage in 3 of 20 intervention clusters in India, 19 of 20 in Benin, and all 20 intervention clusters in Malawi (Fig 2).

**Fig 2.**
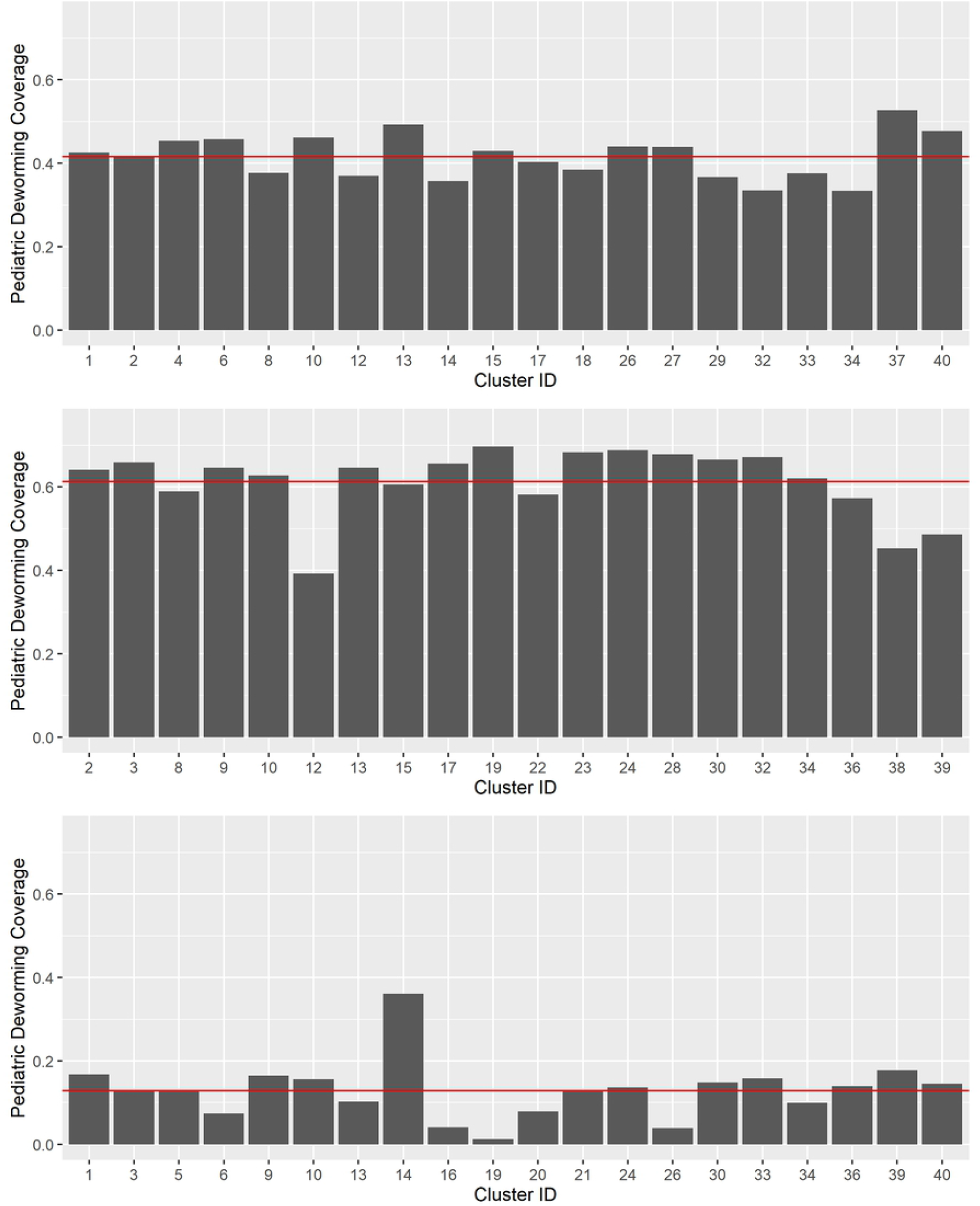
Individual-Level Reporting of SBD in Intervention Clusters, by Study Site. (A) Benin. (B) India. (C) Malawi. Mean SAC Deworming Coverage shown as a red slicer line. Y-Axis set to WHO target of 75% coverage for SAC and PSAC.

These SAC SBD coverages from individual-level data in the intervention clusters were compared to school-level data collected through the standard of care SBD that occurred in all clusters. By comparing the school-level data in the intervention clusters to the gold standard individual-level data in those same clusters, we calculated mean squared errors (MSE) for each cluster which were then averaged to determine a single error value for each site. These errors were 0.096 in Malawi, 0.135 in India, and 0.209 in Benin (Fig 3).

**Fig 3.**
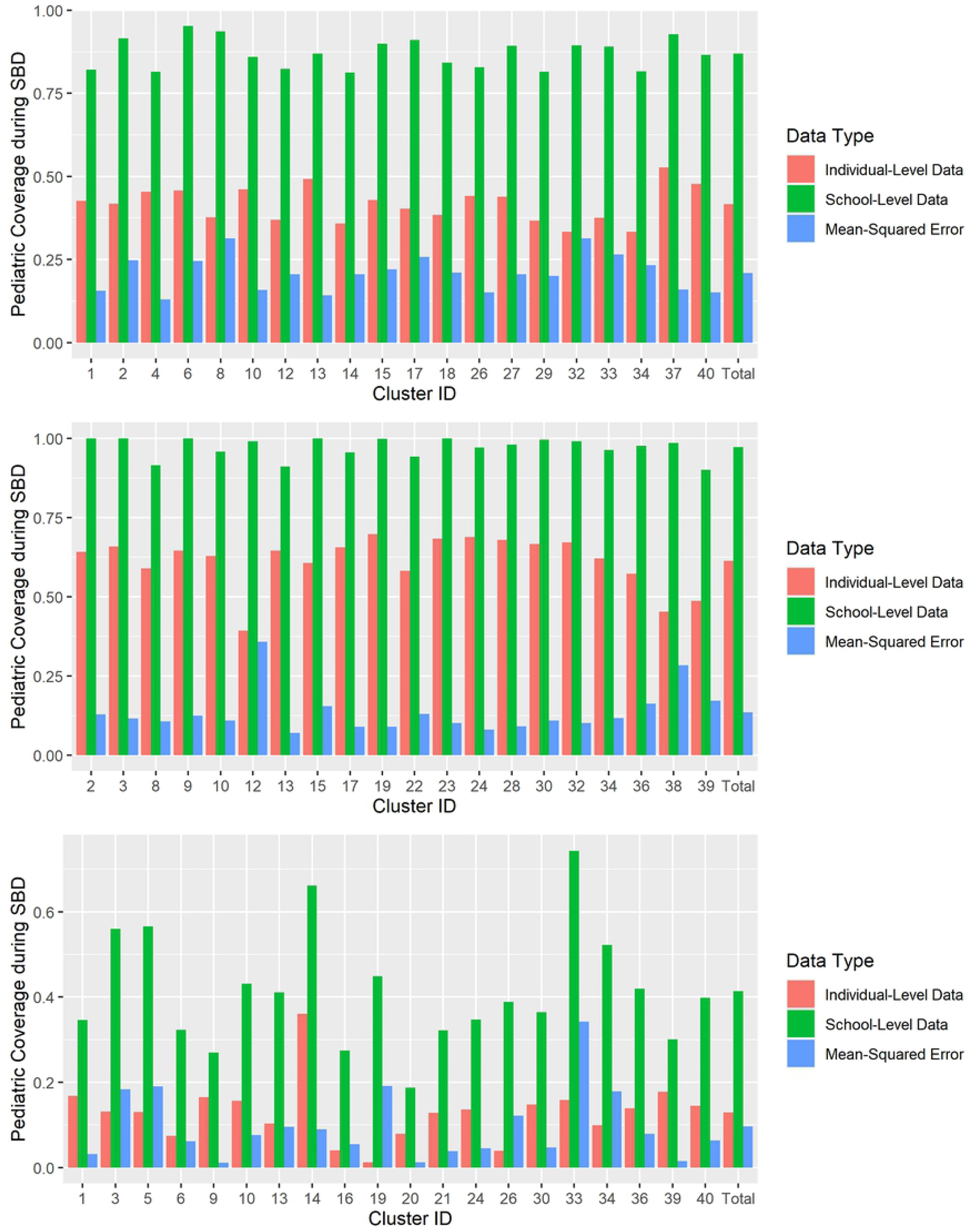
Comparison of cluster-level SAC SBD coverage from individual-level data collection to school-level SBD in intervention clusters, by site. (A) Benin. (B) India. (C) Malawi.

The overall MSE measurements per site were applied to school-level SBD coverages in the control clusters to project individual-level SBD coverage estimates for the control arm of the DeWorm3 trial (Fig 4).

**Fig 4.**
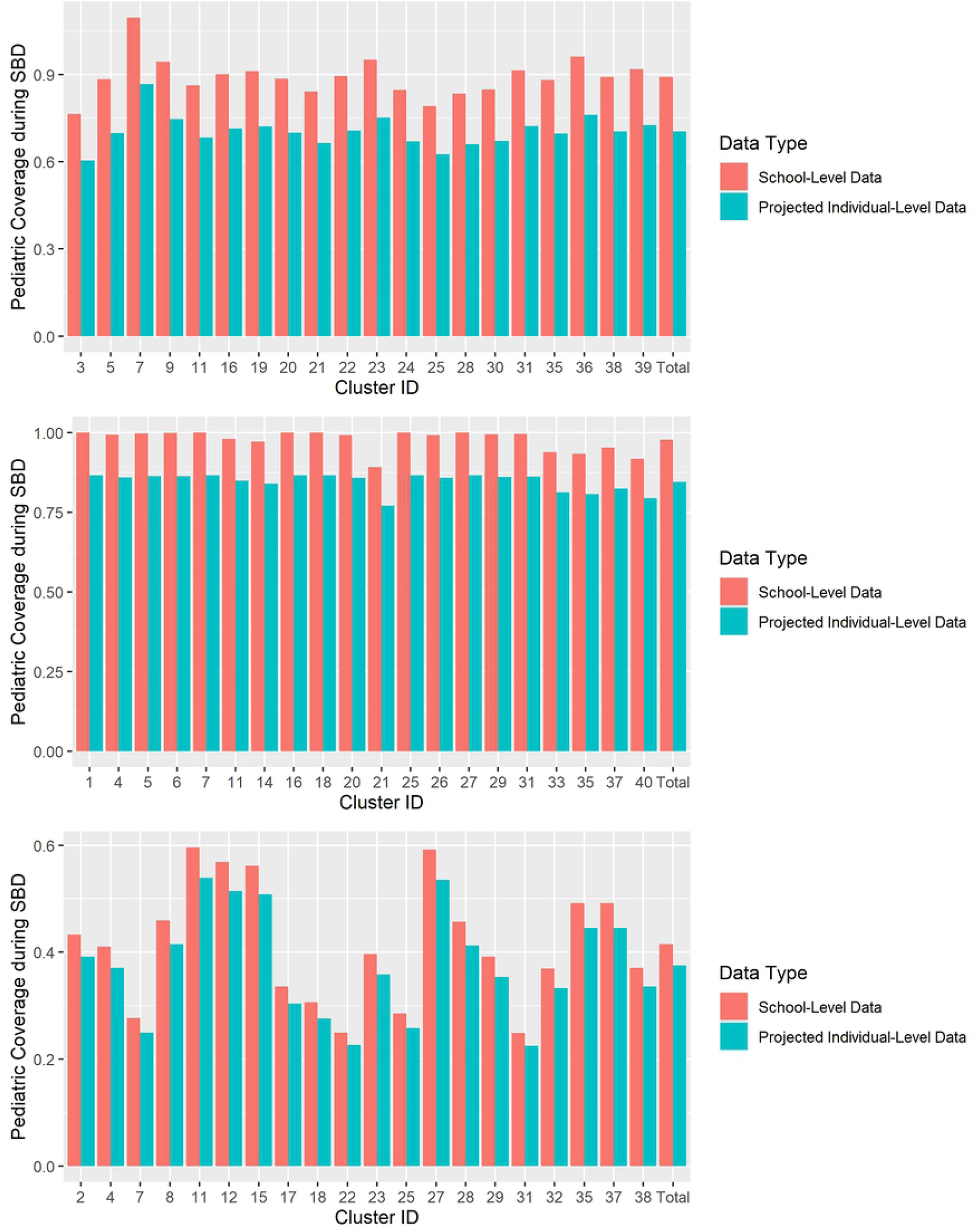
Comparison of school-level SBD coverage estimates to projected SBD coverage estimates in control clusters if individual-level data had been available, by study site. (A) Benin. (B) India. (C) Malawi.

In Benin, school-level SBD data from the trial’s control clusters originally showed an average SAC coverage of 89.1%, compared to 70.5% (p-value < 0.001) when applying observed MSE to project expected coverages if individual-level data had been available. In India, 97.7% SAC school-level SBD coverage for control clusters decreased to 84.5% (p-value < 0.001) when projected for SBD estimates from individual-level data, and in Malawi the SAC school-level SBD coverage decreased from 41.5% to 37.5% (p-value < 0.001) when projected for SBD estimates from individual-level data (Fig 5).

**Fig 5.**
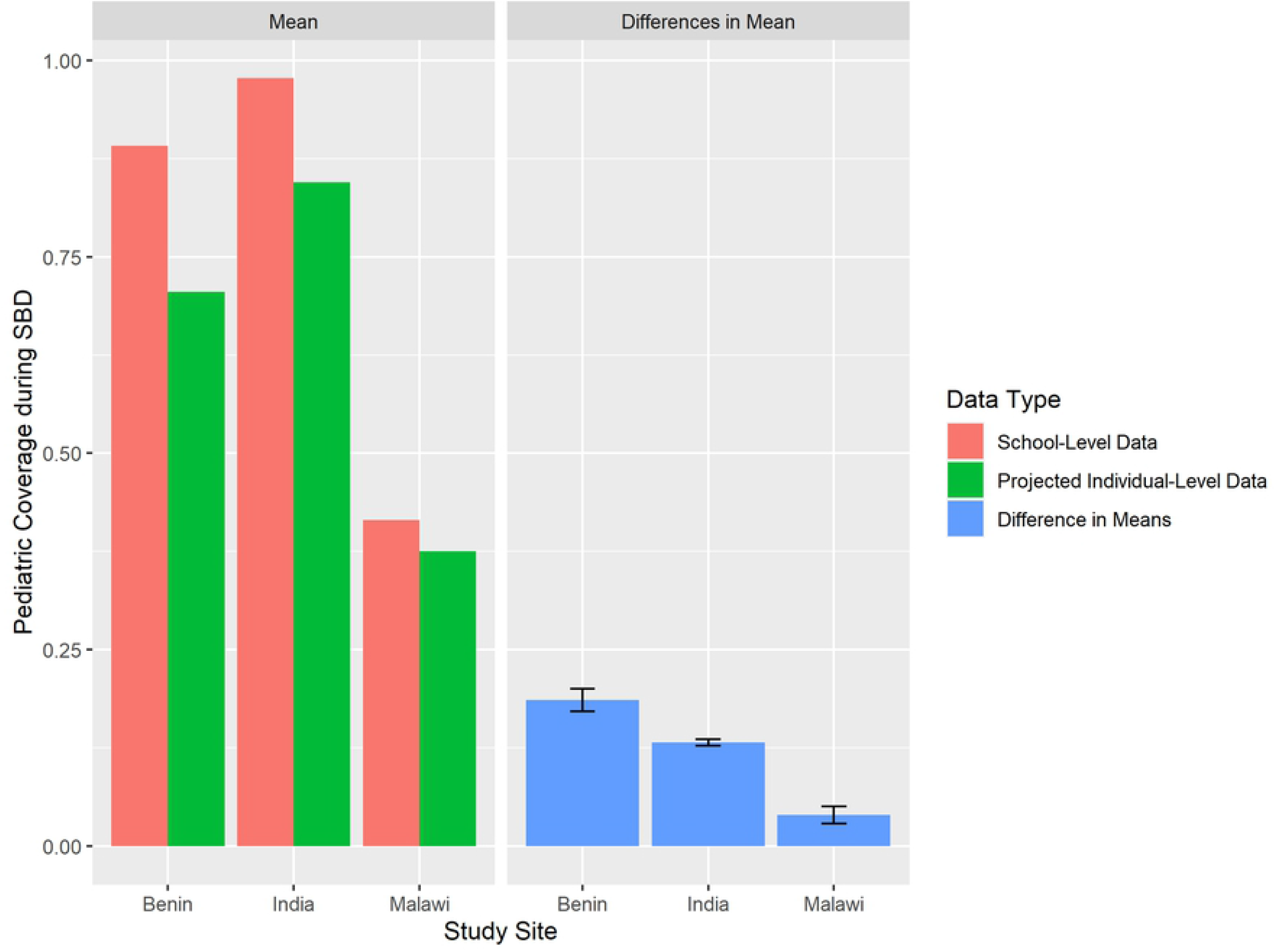
Comparison of school-level average SBD coverage to average individual-level SBD coverage in control clusters after application of mean-squared error rates.

The catchment areas created in this analysis show a high degree of overlap, particularly in the highly populated urban centers of the DeWorm3 study areas. Removal of geographic outliers resulted in fewer instances where catchment areas were greatly extended in order to include a small number of distant SAC. Recalculated school catchment areas were shown to overlap each other less than in the naïve analysis, particularly in Benin (Fig 6A).

**Fig 6.**
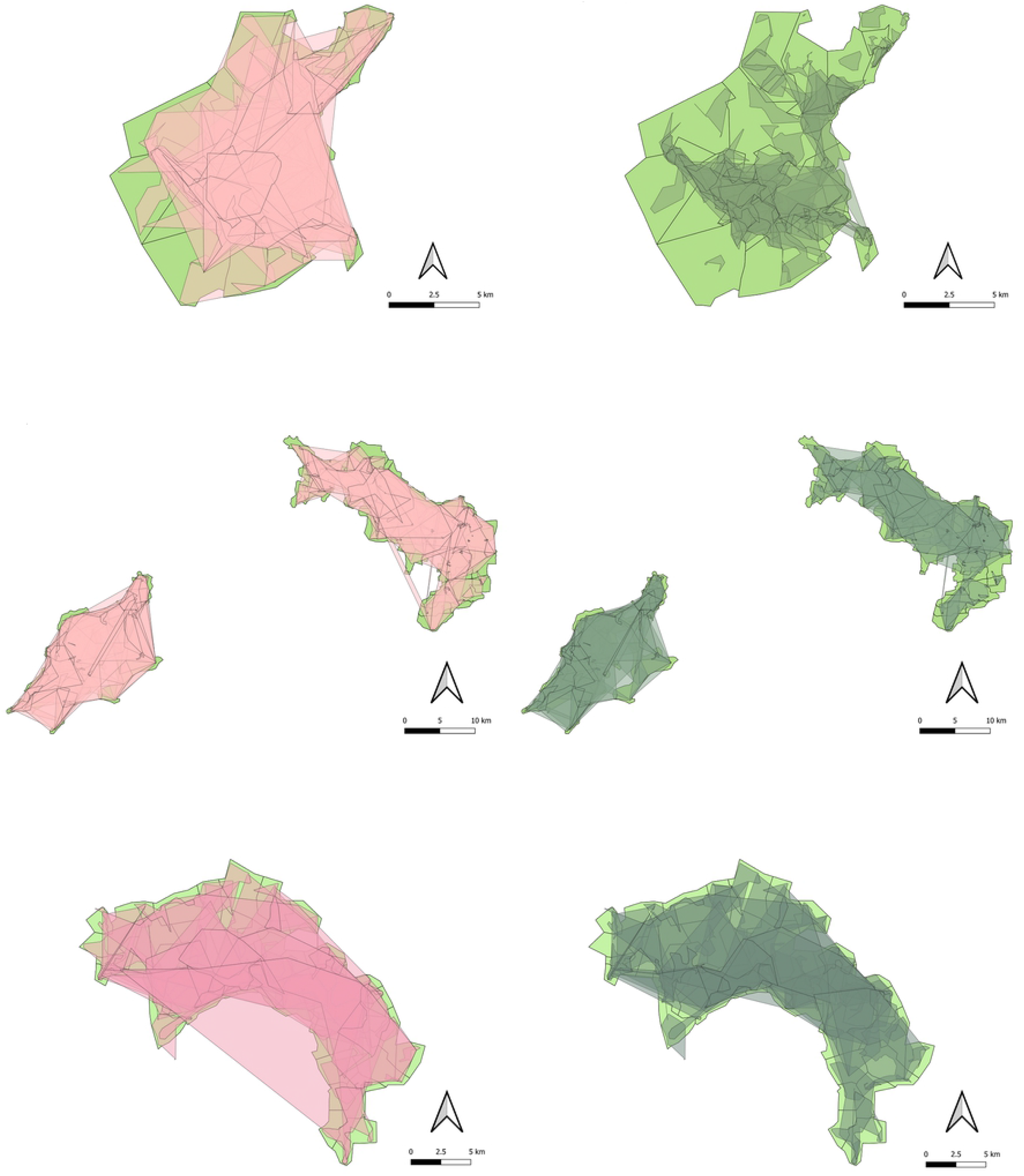
Catchment areas for each school by study area, created via QGIS Concave Hull (K-nearest neighbor) analysis. (A) Benin. (B) India. (C) Malawi. The left side of each panel shows naïve catchment areas in semi-transparent pink created prior to removal of geographic outliers, while the right side shows catchment areas in semi-transparent green created after removal of geographic outliers. Both overlay original cluster boundaries in green.

This reflects the reality that school catchment areas are geographically indistinct and do not merely include the SAC closest to them. This means that any attempt to fix catchment areas solely using geographic proximity is unlikely to be accurate, especially in densely populated urban centers with many schools. For example, of the 40 clusters comprising the Benin study area, in only 7 clusters did more than 50% of SAC attend the school that was closest to their home by Euclidean distance, and in only one cluster was this the case for more than 65% of SAC (Supplemental Table 1). Overall, the percentage of SAC that attended the school geographically closest to their home was 29% in Benin and 10% in India, although that number was substantially higher in Malawi at 66% (Table 2).

**Table 2.** Number of School-Age Children in DeWorm3 Catchment Areas and Proportion Attending the School Geographically Nearest to Their Household, by Study Site.

|  | Number of SAC (n) | Proportion Attending Nearest School |
| --- | --- | --- |
| Benin | 21840 | 0.29 |
| India | 25508 | 0.11 |
| Malawi | 42550 | 0.66 |

## Discussion

The success of global STH programs is critically dependent on the accurate measurement of programmatic treatment coverage. Funding mechanisms, policy directives, and the support of local stakeholders all require that programs can demonstrate the reach and the impact that they provide to local communities. Accurate reporting of data is of fundamental importance to achieving this goal. This paper seeks to enhance our understanding of deworming coverage provided during SBD by demonstrating the error inherent in school-level data collection when compared to a gold standard of individual-level reporting. This paper compares individual-level SBD data collected via community-wide MDA from the DeWorm3 trial’s intervention arm against school-level SBD data collected via school-level reporting in both arms to calculate the difference between these estimates in the intervention arm clusters where both numbers are available. This is a new approach for characterizing discrepancies of SAC deworming coverage during SBD. This analysis was made possible by the DeWorm3 trial’s recording of previous SBD treatment status for SAC reached during community-level MDA.

Cluster-level SBD coverages for SAC derived from individual-level data were substantially lower than SBD estimates derived from school-level reporting, which indicates that school-level SBD reporting may be over-estimating SAC deworming coverage when compared to a gold standard of individual-level reporting. This is an important finding because school-level estimates for SAC deworming are more commonly available to national STH deworming programs than the individual-level data provided in a trial such as DeWorm3. If school-level data are consistently overestimating coverage, then decisions made using these data at the program level may be made under the assumption that SBD coverage is higher than it actually is. Such a discrepancy could result in miscalculation of the duration and frequency of deworming for future planning, premature cessation of MDA or disruptive allocation of resources based on a flawed assumption of treatment coverage.

There may be a number of reasons for the differences between these estimates that are not accounted for by systematic over-estimation of school-level SBD coverage. It is important to note that the weighted average calculation used to determine school-level SBD coverages in this analysis is most heavily influenced by the schools that report the largest number of SAC attendees from school enrollment data in the DeWorm3 trial’s annual school census. If a small number of SAC from a particular cluster report attending a school with low coverage, then that cluster may still have a large number of SAC who attend a nearby school with higher coverage which would decrease the low-coverage signal. This would lead to a possible overestimation of cluster-level coverage when relying solely upon school-level data if higher coverages are associated with larger schools. Additionally, the school-enrollment totals available during SBD may have included SAC that do not usually attend that school but who showed up for SBD days. This may have been the case in Benin and India, where overall enrollment totals on the day of SBD exceeded enrollment totals in the trial’s school census, as opposed to in Malawi, where there were fewer students listed as enrolled on the day of deworming than in the school census. If these SAC who are arriving for deworming treatment days but are not regularly enrolled are more difficult to reach during community-wide MDA data collection, then individual-level reporting in the intervention arm may be underestimating true treatment coverage of SBD, leading to an erroneously high depiction of over-estimation in the school-level SBD data.

The finding that coverage reported by schools or district health authorities is consistently higher than coverage as determined by individual-level survey responses has been previously observed in trials (16, 17, 18), and when comparing national reporting of school-based treatment to WHO versus individual-level Demographic and Health Survey results (19). Previous papers have postulated that the use of school or district-level reported coverages may open studies to biased results that overestimate deworming coverage due to exaggeration by community drug distributors, local health officials, or schoolteachers (6). Another standard concern for survey-based data collection is recall bias, although this is unlikely to have affected the low coverages reported during individual-level data collection, as the DeWorm3 trial utilized indelible ink to mark the fingers of SAC who were treated at school in order to mitigate this concern (20). In discussions with DeWorm3 program staff, it was noted that SBD coverages derived from individual-level data collection for the data analyzed in this paper might have been unusually low in Malawi because of a longer-than-usual gap between SBD and community-wide drug distribution during that year, which would normally occur within two weeks. It is possible that this may have resulted in lower estimates for previous treatment during SBD because the ink may have faded from the fingers of SAC, preventing the use of this marking for accurate measurement of previous SBD treatment.

This paper also utilized concave-hull geospatial analysis to define the catchment areas of schools in the DeWorm3 trial areas for the purposes of estimating SAC deworming treatment coverage. Previous approaches have relied upon proximity-based (21), Bayesian modeling (22), and cost-distance approaches (23) to estimate likely catchment areas for service delivery of health posts, hospitals, and schools for deworming activities. The DeWorm3 trial provides access to geolocation information for each SAC treated within the study areas, allowing for exact visualization of reported catchment areas. The school catchment areas in the DeWorm3 trial areas overlap each other significantly, especially in the population centers where schools are geographically near to one another. In addition, most SAC in these study areas did not receive their deworming treatments at the school geographically nearest to them. If this is the case for populations in other study areas, this means that proximity-based approaches for defining health activity catchment areas may introduce considerable inaccuracies for determining relevant population denominators. A lack of clear communication to survey administrators regarding the geographic limits of clusters, municipal boundaries, and catchment areas has been previously identified as a possible driver of variation in deworming treatment coverage (24, 6). Therefore, a potential benefit stemming from the visualization of these catchment areas may include more streamlined data collection and drug-distribution practices for school-based and community-wide MDA in the DeWorm3 trial area.

The DeWorm3 trial is an exceptionally large and rigorously conducted study, with harmonized data collection practices across three different countries. Additionally, these data had already gone through initial data cleaning from the study sites and the central DeWorm3 data team, resulting in overall uniformity and confidence for use in analysis.

However, there were a number of limitations that should be considered when interpreting these findings. The uneven quality of self-reported school attendance data used to create catchment areas and to assign population-based weights to each school in the cluster-level weighted average coverage calculations may have impacted these results. In the case of catchment-area creation, naïve catchments from the self-reported data were modified to exclude wide-ranging geographic outliers to more accurately predict where a hypothetical child for whom self-reported school attendance is unavailable would likely go for deworming treatments in a given cluster. And for the assignment of population-based weights, the self-reported attendance data do not allow for consideration of those SAC who may not have been reached during data collection, or whose responses were matched to the incorrect school.

Free-text school name entries also pose a limitation because there is no perfect recording of where each child in a given cluster received their SBD treatment. To minimize the effect of this limitation, thousands of free-text entries were analyzed and matched, where possible, to the appropriate school in each study site’s school registry, via the iterative process described in the methods section. In this analysis, 17% of SAC respondents in Benin were not matched to a specific school, while 15% and 10% of SAC remained unmatched to a school in India and Malawi, respectively. These unmatched SAC reported going to schools that were unknown, were outside the study area, or were schools for which coverage levels and enrollment data were not available, with informal or nursery schools accounting for the largest percentage of free-text entries that cannot be matched to a registered school. As an illustrative example, it is believed that over 97% of the 3,929 unmatched free text entries for the India study site of the DeWorm3 trial refer to Anganwadi (informal government-run education centers for rural childcare developed under the Integrated Child Development Scheme) centers due to similarities in the submitted free-text responses. This has been a limitation in previous studies, including in Bangladesh, where informal schooling complicated calculation of coverage and population statistics (25). If large numbers of these unmatched respondents actually attended one of the schools in the school registry, then the weighted average formula for determining coverage in SBD from school-level data would have slightly underweighted the schools that they attended. This would only provide a significant limitation if large numbers of SAC were assumed to attend an erroneous school in the registry, leading to a slight overweighting of that school in the weighted average coverage formula.

Finally, it is important to note that this analysis only utilized data collected in year two of the DeWorm3 trial, which consisted of two treatment rounds in India and 1-2 rounds in Benin and Malawi, as these data were determined by the DeWorm3 program staff to be the most complete dataset available. In year one, data were less reliable because certain data collection processes had not yet been standardized, and in year three, data collection was disrupted by the worldwide emergence of SARS-CoV-2. A more complete analysis would examine data across multiple study years to determine if SAC coverage rates fluctuated significantly from year to year, and if observed error in the school-level SBD data were sustained throughout the duration of the trial.

## Conclusion

This analysis utilized data from the DeWorm3 trial to quantify discrepancies between cluster-level estimates of SAC deworming coverage derived from gold-standard individual-level data collection during community-wide MDA in the intervention arm and historically less reliable school-level data collection during the delivery of standard of care school-based deworming in both trial arms. These estimates, derived from three different country sites and across many settings within those countries, indicate that school-level reporting of SAC deworming coverage likely overestimate program coverage in these age groups. These results suggest that school-level reporting of SAC deworming coverage should be validated by individual-level data collection. Novel interventions to improve data collection within MDA programs are needed to ensure that accurate reporting informs programmatic decision-making and resource allocation.

## Data Availability

Data cannot be shared publicly because the study remains blinded to outcome data. Data are available from the DeWorm3 Institutional Data Access Committee (contact via Barbra Richardson) for researchers who meet the criteria for access to these data.

## Acknowledgements

The authors wish to thank all of the study participants, communities and community leaders, national NTD program staff and other local, regional and national partners who have participated or supported the DeWorm3 study. We also would like to acknowledge the work of all members of the DeWorm3 study teams and affiliated institutions. We give special thanks to the Clinical Trial and Implementation Science Support Unit at the University of Washington for their aid in the conception of this analysis.

## Supporting Information

**S1 Table. Cluster-level School Age Children Statistics, by Study Site** Cluster-level statistics of the proportion of SAC attending the school that is geographically closest to their home, the total number of students, and the number of unique schools reported as being attended by students in each cluster, replicated for each of the three study sites.

